# The changing major trauma disease burden from low falls in the first two decades of the 21st Century – a longitudinal analysis from the largest European Trauma Registry

**DOI:** 10.1101/2022.05.16.22275136

**Authors:** Thomas AG Shanahan, Michael Tonkins, Omar Bouamra, Dhushy Surendra Kumar, Antoinette Edwards, Laura White, Anthony Kehoe, Jason E Smith, Timothy J Coats, Fiona Lecky

## Abstract

**Objective:** The 21st century has seen an unexpected rise in numbers of patients with severe injuries caused by low energy transfer mechanisms such as low falls. Our objective was to determine whether this increasing “low energy transfer major trauma” disease burden is more strongly associated with population ageing, better detection or registry reporting between 2000 and 2019.

**Design:** Longitudinal series of annual cross-sectional studies from the Trauma Audit Research Network (TARN).

**Setting:** England and Wales.

**Participants:** Patients with an Injury Severity score (ISS) >15, admitted to English or Welsh hospitals from January 2000 until December 2019.

**Main outcome measures:** The primary outcome was the strength of association of changes in annual rates of; major trauma patients receiving three-dimensional imaging (CT/MRI scans), registry reporting, and proportion of the population aged over seventy-five with changes in the annual proportion of patients injured via low energy transfer mechanisms.

**Results:** The annual proportion of major trauma patients injured by low energy transfer mechanisms rose more than four-fold from 12.5% in 2000 to 52.6% in 2019 (+40.1%, 95% CI 38.8 to 41.4, p<0.0001). This rise in the prevalence of low energy major trauma was more strongly associated with case ascertainment improvements over the study period - indicated by a 60% increase in the proportion of major trauma patients receiving CT/MRI (+60%), and 47% increase in registry reporting rates than a one percent rise in the prevalence of people aged 75 years and over in England and Wales.

**Conclusion:** Between 2000 and 2019 changes in major trauma imaging and reporting have revealed the previously hidden burden of injury resulting from low energy transfer mechanisms, potentially explaining significant increases in major trauma patient numbers. Future research should recognise low energy and high energy major trauma patients are distinct groups and require different interventions to improve patient outcomes.

**Section 1: What is already known on this topic:**

- Falls from standing height or less than two metres are causing an increasing major injury disease burden across Europe.
- Internationally trauma systems have focussed on patients with injuries caused by high energy transfer mechanisms, such as road traffic collisions.

**Section 2: What this study adds:**

- Better detection and reporting of low energy transfer major trauma - rather than population ageing - appear to be are driving the increase in lower energy major trauma.
- Low energy transfer major trauma - characterised by older age, less abnormal physiology, but a high prevalence of traumatic brain and thoracic injuries - is now the dominant major trauma (ISS>15) presentation in England and Wales.
- Low energy transfer major trauma patients wait longer for CT scan, are less likely to receive the care in the highest-level trauma centre, to undergo surgery or be admitted to critical care.

**Section 3: How this study might affect research, policy and practice:**

- Future research should recognise high energy and low energy trauma patients are distinct groups, which require different interventions to improve care processes and outcomes.

## Introduction

Globally injuries sustained through high energy transfer mechanisms, such as road traffic collisions (RTCs) are ranked as the top cause of disability-adjusted life-years (DALYs) lost for people aged 10-49 years (1). However, recent studies in high income countries have identified injuries from low energy transfer mechanisms - falls from a standing height or less than two metres - as the most prevalent cause of major trauma presenting to hospital. Low energy major trauma occurs predominately in older people (>64 years) (2–4) who are more likely to fall and to sustain life threatening injuries due to reduced physiological reserve.

Previous studies have identified three potential drivers of the increase in low energy transfer major trauma over time. First, it is suggested the rise is due to an ageing population(2). Second, a change in imaging practice has been identified as a driver.(5) The use of three dimensional imaging (computed tomography or magnetic resonance scans) has increased significantly in the 21^st^ century, and is now advocated as a first line diagnostic modality in preference to plain x-rays.(6) Third, membership of and better reporting practices to trauma registries have been posited as a driver for the increase in low energy transfer major trauma. For their part, trauma registries have become integrated into trauma care system quality assurance and improvement with compulsory reporting(7). Both are likely to create changes in major injury case ascertainment particularly in low energy major trauma. However, these drivers have not been simultaneously examined in an inclusive trauma system or using data beyond 2013.

An accurate understanding of the aetiology and epidemiology of major trauma is necessary for effective injury prevention and optimisation of trauma care systems – both of which currently prioritise resources for those injured by high energy transfer mechanisms. Therefore, the primary aim of this study was to determine the temporal associations between the changing prevalence of low energy transfer as the causal major injury mechanism and changes in imaging practice, trauma registry reporting and population age in England and Wales in the 21^st^ century. The secondary aim was to assess associated temporal trends in major trauma patient volume, characteristics, care pathways and outcomes over the study period informed by a comparative cohort assessment of patients injured by low and high energy transfer mechanisms.

## Methods

### Study design and setting

This is a longitudinal series of annual cross-sectional studies from the Trauma Audit Research Network (TARN – the national clinical audit for major trauma care) using a consistent methodology. The study design and reporting followed the STROBE guidance for observational studies(8). For this study, anonymised data from major trauma patients presenting to English and Welsh hospitals between 1st January 2000 and 31st December 2019 were collated for analysis. The UK Health Research Authority Patient Information Advisory Group (PIAG) has given approval (Section 20) for TARN analysis of anonymised patient data. Major trauma was defined as a patient with an Injury Severity Score (ISS) >15.

TARN includes patients of all ages who sustain injury resulting in hospital admission >three days, critical care admission, transfer to a tertiary/specialist centre or death within 30 days of hospital arrival. Isolated femoral neck or single pubic ramus fracture in patients >65 years and simple isolated injuries are excluded. After study inclusion, a dataset of prospectively recorded variables covering demographics plus injury-related physiological, investigation, treatment and outcome parameters are collated using a standard web-based case record form by trained TARN hospital audit co-ordinators. Injury descriptions from imaging, operative and necropsy reports are submitted by TARN co-ordinators. All injuries are coded centrally using the Abbreviated Injury Scale (AIS), which enables calculation of the Injury Severity Score (ISS). The ISS is used to assess the overall severity of a patient’s injuries by weighting the severity of injuries to each of six body regions(9).

For the primary analysis, mechanism of injury for each patient was categorised as either ‘high energy’ or ‘low energy.’ Low energy mechanisms comprised falls from two metres or less including ground level falls. High energy mechanisms comprised all other mechanisms (e.g., road traffic collisions, falls at or > two metres, sport associated injuries and assault/self-harm).

### Primary Outcome

The outcome (dependent variable) used in the statistical analysis was the strength of association between the temporal trend in the annual proportion of major trauma sustained through low energy transfer mechanisms and putative causal or associated factors. We contemporaneously assessed (a) the annual percentage of study patients receiving CT or MR imaging, (b) the annual percentage of patients with TARN-eligible international classification of disease (ICD) injury codes in hospital episode statistics (HES - denominator) entered onto the TARN database and (c) mid-year estimates of the proportion of the English and Welsh population aged over 75 (Office of National Statistics) ONS. HES data was obtained from NHS Digital and ONS reports via the Nomis service.

The secondary analysis described the changes over time in annual major trauma patient numbers, characteristics, and care. Patient age, sex, comorbidities, anatomical distribution, severity of injury and presenting physiology were the characteristics examined. Any Abbreviated Injury Score (AIS) 3+ severity injury was defined as significant in a specific body region; polytrauma was defined as >one body region with an AIS 3+ injury. We similarly assessed care from prehospital medical teams, administration of tranexamic acid (TXA), consultant care in the emergency department, endotracheal intubation, surgery within 24 hours of first hospital arrival, critical care admission and highest level of hospital trauma accreditation - a Major Trauma Centre (MTC), the highest level in the National Health Service – as either the first receiving hospital or during the acute care pathway. The second level of care in a Trauma Unit (TU) reflects a trauma receiving hospital accredited for resuscitation, general admissions, and secondary transfer to an MTC.

We also report time from hospital arrival to initiation of the first CT and to first operation within 24 hours. The patient outcomes assessed included length of acute care stay, length of stay in critical care and acute care hospital mortality.

To understand the impact of temporal changes in causal mechanism a further secondary analysis compared the above characteristics by energy transfer level in major trauma patients presenting in 2019. We closed the study at the end of 2019 as the impact of the COVID-19 lockdown on the major trauma disease burden and patient outcomes requires an ongoing separate evaluation.

### Analysis

The study size was determined by the number of eligible patients included in the TARN database. The prevalence of major trauma resulting from low energy transfer, major trauma patients receiving CT or MRI, case ascertainment, proportion of England and Wales population aged seventy-five and over, patient demographic features, injury characteristics, patterns of care and mortality were described by year of presentation. To define the changes that took place, trends in annual proportion of patients injured by low energy transfer, case ascertainment, imaging, population proportion over 75 years and secondary outcome categorical variables were examined using Chi-squared test for trend. A two-sided p value of < 0.05 was statistically significant. As small changes within such a large dataset give rise to ‘statistical significance,’ only changes that were considered to also have ‘clinical significance’ were analysed for trend or highlighted in the text. All data were presented in the tables which report 2019 v 2000 t-tests on difference of proportion for categorical variables and the Bonnet-Price Test on difference of medians for continuous variables(10).

A statistical model to assess the primary outcome was derived. Because of the non-linear nature of the independent variable (percentage injured by low energy transfer) a weighted generalised linear model with logit link and binomial family was used with robust standard error to correct for any model misspecification. This was followed by marginal effect analysis to identify factor(s) whose change was most strongly associated with the primary outcome.

In the analysis of hospital mortality, mortality at 30 days or at discharge (whichever occurred first) was used as the dependent variable, with admission year as an independent exposure variable, to enable the calculation of odds of death for each year from 2001 to 2019 compared to the baseline year 2000. The case-mix adjustment of the logistic regression model used Glasgow Comma Score (GCS), ISS, age, an age/gender interaction, and the modified Charlson Comorbidity Index(11). The GCS was not recorded in 19811 (8.2%) of the selected cases, and to include them in the regression model, an imputation technique based on chained equations and Rubin’s rules, was therefore used on the assumption that the mechanism of missingness is at random (MAR) that is the missing value depends on measured variables. The imputation model contained all the variables included in the regression model and the outcome(12). The analysis was performed with the software version 16.1 (StataCorp. 2019. *Stata Statistical Software: Release 16*. College Station, TX: StataCorp LLC).

To adjust for selection bias arising from increasing hospital membership of TARN over the study period, a sensitivity analysis assessed the primary and secondary outcomes in a subgroup of 19 large hospitals consistently submitting to TARN (2000-2019).

## Results

A total of 241,484 patients were included in the primary analysis, of which 96,833 (40%) were injured by low energy trauma and 144,651 (60%) were injured by high energy transfer mechanisms (Figure 1).

**Figure 1:**
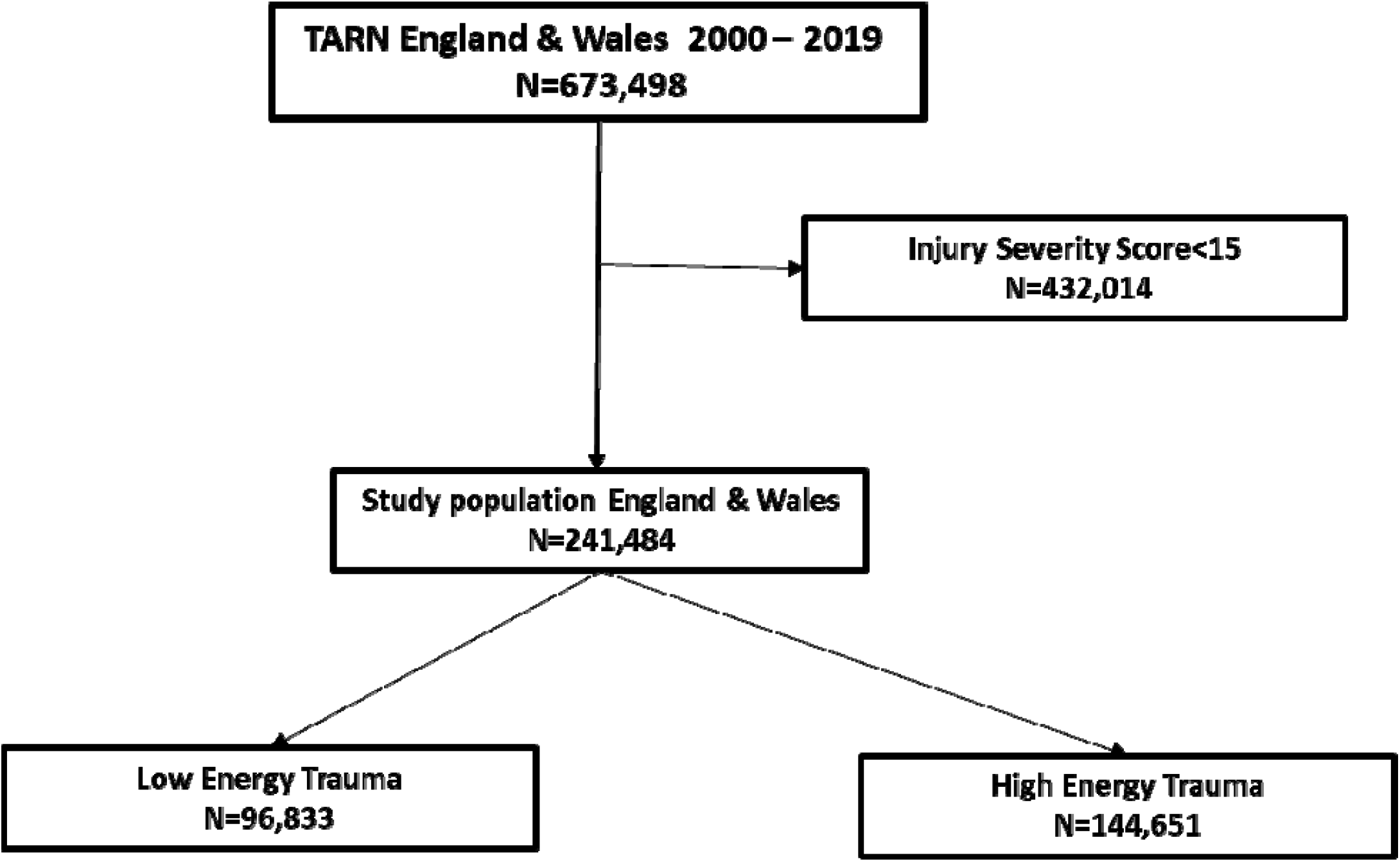
Identification of eligible major trauma patients from TARN with Injury Severity Score >15; classified by energy transfer mechanism. Low energy = falls from standing or from height < 2 metres. High energy trauma was defined as road traffic collisions, falls from height of 2+ metres, sporting injuries, assault, stabbing and shootings, (Trauma Audit and Research Network = TARN).

### Temporal changes in proportion of major trauma caused by falls from standing or less than two metres and associated factors

There has been a significant temporal trend (chi squared p<0.001) and four-fold increase in the proportion of major trauma sustained through low energy mechanisms. Low-energy transfer mechanisms accounted for 12.5% of major trauma patients in 2000 (n = 373), rising to 52.6% (n = 16,087) in 2019 (Table 1), with most of the increase in prevalence (320% in relative terms) occurring by 2017 (Figure 2 chi squared test for trend p<0.001).

**Table 1:**
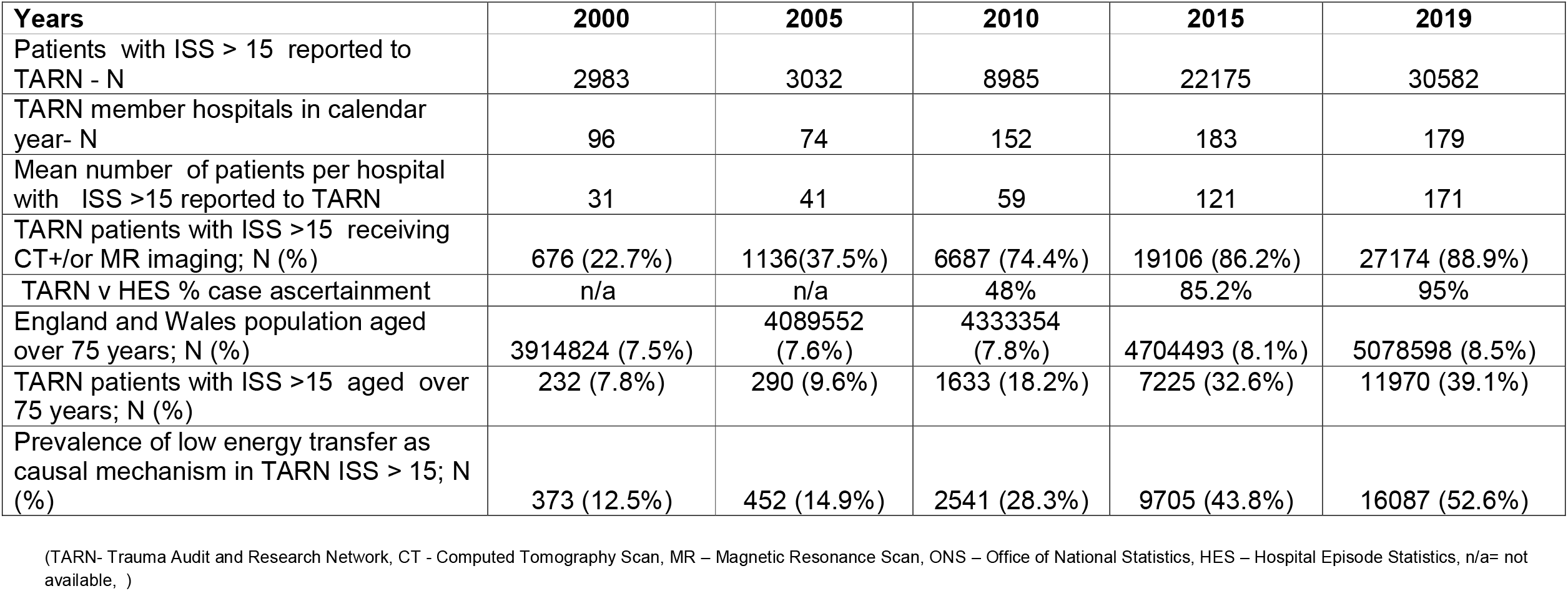
Annual prevalence of patients injured by low energy major trauma (primary outcome) and factors assessed for association in English and Welsh hospitals submitting to TARN in 2000, 2005, 2010, 2015, 2019.

**Figure 2:**
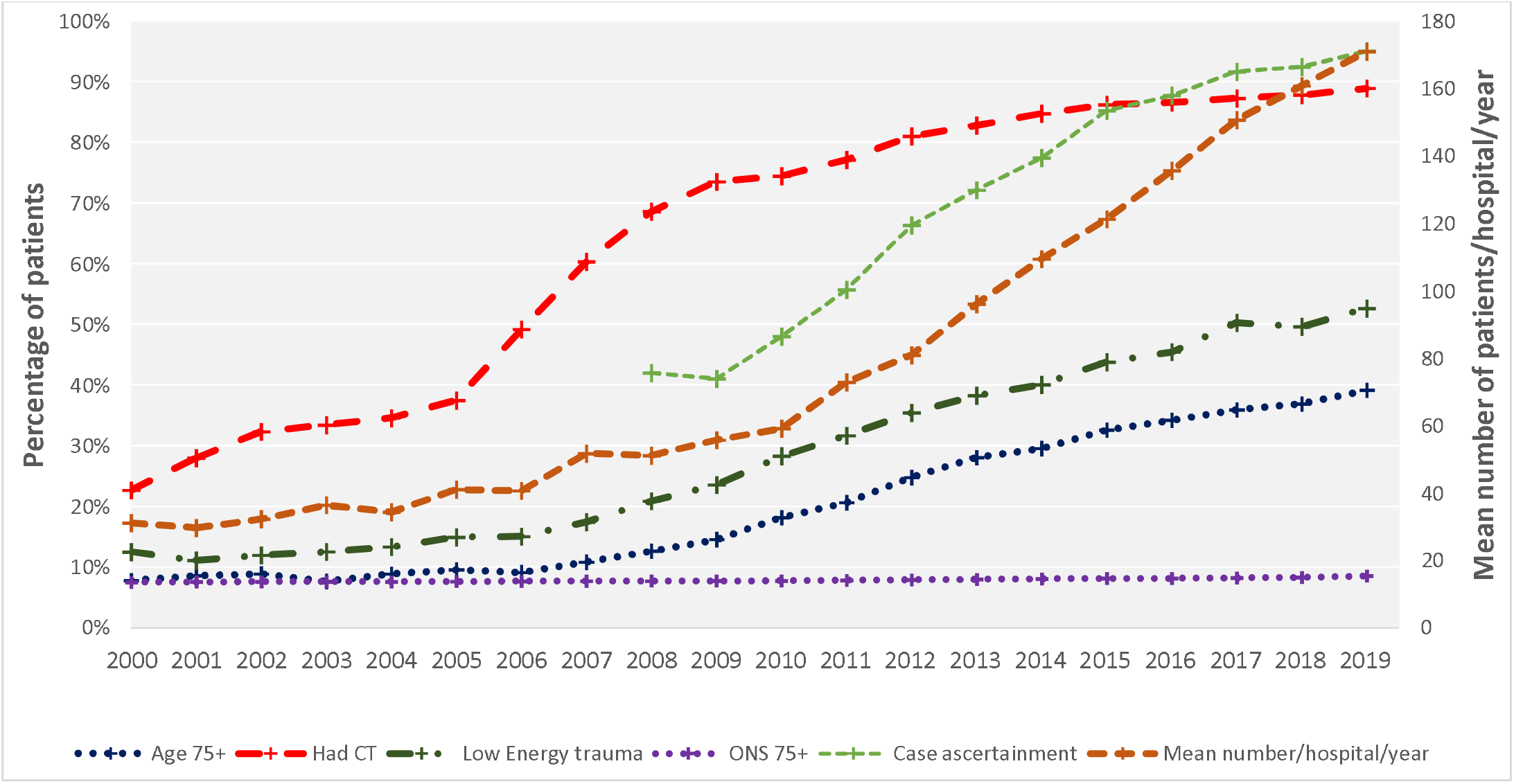
Contemporaneous annual (2000-2019) percentage of TARN ISS>15 patients injured through low energy transfer and factors assessed for temporal association; percentage of TARN (i) aged 75 years and over (ii) receiving CT or MR scanning; percentage of population of England and Wales aged 75 years and over, mean number of ISS>15 patients reported per hospital to TARN, percentage TARN case ascertainment (denominator = TARN eligibility in Hospital Episode Statistics 2008-2019). (TARN-Trauma Audit and Research Network, CT - Computed Tomography Scan, MR – Magnetic Resonance Scan, ONS – Office of National Statistics, HES – Hospital Episode Statistics)

The proportion of major trauma patients receiving CT or MR imaging has increased. In 2000, 676 major trauma patients (22.7%) underwent CT or MR, whereas by 2019 this had risen to 27,174 (88.9%) (Table 1). Most of this change had occurred by 2012 (Figure 2; chi squared test for trend p<0.001).

TARN hospital membership within England and Wales doubled between 2000-2019 from 96 to 179 hospitals. Over the same period the annual mean number of major trauma cases per hospital per annum increased almost six-fold from 31 to 171 (Table 1 and Figure 2). Case ascertainment increased over the study period from 42% in 2008 to 95% in 2019 (Table 1); most of this increase had occurred by 2016 (Figure 2; chi squared test for trend p<0.001).

During the study period the population of England and Wales increased by 14% from 52,140,181 in 2000 to 59,439,840 in 2019. Over the same period the number of individuals aged over 75 years increased by 29.7% from 3,914,824 to 5,078,598. Therefore in 2019, individuals aged >75 years accounted for approximately 8.5% of the total population of England and Wales (Table 1) when compared to 7.5% in 2000 – a 1% absolute and 13% relative increase (chi squared test for trend p<0.001).

The analysis of marginal effects demonstrated the relative percentage increase in the annual proportion of low energy major trauma patients associated with 1% increases in annual rates of; CT/MR imaging, case ascertainment versus HES and population prevalence of people aged seventy-five and over was 35.8%, 34.2% and 17.5% respectively.

The sensitivity analysis performed using those hospitals that consistently submitted data during the study period revealed comparable results. There was an almost three-fold increase in the prevalence of major trauma patients injured by low energy transfer mechanisms from 15.3% (n=1790) in 2000 to 43.1% (n=3263) in 2019, associated with increasing rates of imaging, reporting to TARN and overall patient numbers (suppl Table 1, suppl Figure 2; chi squared test for trend p<0.001).

### Temporal changes in overall major trauma patient cohort

The temporal analysis of major trauma patient demographics, injury patterns, presenting physiology and care pathways are reported in Table 2, Figure 2, and Figure 3. The median age of major trauma patients increased significantly from 35.2 to 67.3 years. This is compared to an increase in the England and Wales population median age from 37.9 years in 2001 to 40.2 years in 2019 (chi squared test for trend p<0.001). The proportion of patients aged >75 years in TARN increased five-fold from 7.8% to 39.1%, whilst in England and Wales the proportion aged >75 increased from 7.5% in 2000 to 8.5% in 2019. The proportion of major trauma patients of male sex fell from 74.6% to 63.3%, whilst the proportion of patients with any co-morbidity rose from 15.8 to 74.2% (Figure 3; chi squared tests for trend both p<0.001).

**Table 2:** Annual Major Trauma Patient Cohort Characteristics: case mix, care pathway and acute care outcomes in 2000, 2005, 2010, 2015, 2019: All hospitals submitting to TARN

|  | 2000 | 2005 | 2010 | 2015 | 2019 | Total 00–19 | Magnitude of % change | P-value magnitude of % change* |
| --- | --- | --- | --- | --- | --- | --- | --- | --- |
| **Demographics** |  |  |  |  |  |  |  |  |
| Male, n (%) | 2225 (74.6%) | 2239 (73.8%) | 6330 (70.5%) | 14688 (66.2%) | 19359 (63.3%) | 161849 (67%) | -11.3 (-12.9 to -9.6) | <0.0001 |
| Age, median (IQR) | 35.2 (20.8 – 53.9) | 37.6 (21.6 – 56.7) | 46.8 (26.6–7.8) | 60.5 (36.7 – 79.8) | 67.3 (43.8 – 82.5) | 56.4 (33.1–78) | 32.1 (30.9 – 33.2) | <0.0001 |
| 0 - 24 yrs, n (%) | 953 (31.9%) | 955 (31.5%) | 2010 (22.4%) | 3135 (14.1%) | 3275 (10.7%) | 39984 (16.6%) | -21.2 (-22.9 to -19.5) | <0.0001 |
| 25 - 49 yrs, n (%) | 1129 (37.8%) | 1079 (35.6%) | 2860 (31.8%) | 5211 (23.5%) | 6022 (19.7%) | 62198 (25.8%) | -18.2 (-19.9 to -16.3) | <0.0001 |
| 50 - 74 yrs, n (%) | 669 (22.4%) | 708 (23.4%) | 2482 (27.6%) | 6604 (29.8%) | 9315 (30.5%) | 69151 (28.6%) | 8 (6.4 to 9.6) | <0.0001 |
| 75 yrs and above, n (%) | 232 (7.8%) | 290 (9.6%) | 1633 (18.2%) | 7225 (32.6%) | 11970 (39.1%) | 70151 (29%) | 31.3 (30.2 to 32.5) | <0.0001 |
| Pre-existing medical conditions, n (%) | 472 (15.8%) | 1067 (35.2%) | 5133 (57.1%) | 14952 (67.4%) | 22685 (74.2%) | 153025(63.4%) | 58.3 (56.9 to 59.7) | <0.0001 |
| **Mechanism** |  |  |  |  |  |  |  |  |
| High energy trauma n(%) | 2610 (87.5%) | 2580 (85.1%) | 6444 (71.7%) | 12470 (56.2%) | 14495 (47.4%) | 144651 (59.9%) | -40 (-41.4 to -38.8%) | <0.0001 |
| Low energy trauma n(%) | 373 (12.5%) | 452 (14.9%) | 2541 (28.3%) | 9705 (43.8%) | 16087 (52.6%) | 96833 (40.1%) | 40 (38.8 to 41.4) | <0.0001 |
| **Vital signs** |  |  |  |  |  |  |  |  |
| Mild GCS 13-15 n(%) | 1466 (49.1%) | 1510 (49.8%) | 4897 (54.5%) | 16446 (74.2%) | 24209 (79.2%) | 167830 (69.5%) | 30 (28 to 38.9) | <0.0001 |
| Moderate GCS 9-12 n(%) | 361 (12.1%) | 275 (9.1%) | 819 (9.1%) | 1755 (7.9%) | 2049 (6.7%) | 19151 (7.9%) | -5.4 (-6.6 to -4.2)- | <0.0001 |
| Severe GCS 3-8 n(%) | 934 (31.3%) | 724 (23.9%) | 1298 (14.4%) | 3025 (13.6%) | 3195 (10.4%) | 34692 (14.4%) | -20.8 (-22.6 to -19.2) | <0.0001 |
| Not recorded n(%) | 222 (7.4%) | 523 (17.2%) | 1971 (21.9%) | 949 (4.3%) | 1129 (3.7%) | 19811 (8.2%) | -3.7 (-4.7 to -2.8) | <0.0001 |
| GCS on arrival, median (IQR) | 13 (6 - 15) | 14 (7 - 15) | 14 (11 - 15) | 15 (13 - 15) | 15 (14 - 15) | 15 (13 - 15) | 2 (2 to 2) | 0.999 |
| SBP mmHg on arrival, median (IQR) | 130 (112 - 150) | 130 (112 - 149) | 134 (117 - 153) | 135 (118 - 155) | 136 (119 - 156) | 135 (117 - 154) | 6 (5.5 to 6.5) | <0.0001 |
| % O2Sats on arrival, median (IQR) | 98 (96 - 100) | 99 (96 - 100) | 98 (96 - 100) | 98 (96 - 100) | 98 (96 - 99) | 98 (96 - 100) | 0 | <0.999 |
| Heart rate on arrival bpm, median (IQR) | 90 (75 - 110) | 88 (73 - 107) | 85 (72 - 102) | 83 (71 - 98) | 82 (70 - 96) | 84 (71 - 99) | -8 (-9 to -7) | <0.0001 |
| **Injuries** |  |  |  |  |  |  |  |  |
| Penetrating | 97 (3.3%) | 126 (4.2%) | 323 (3.6%) | 504 (2.3%) | 655 (2.1%) | 6488 (2.7%) | -1.1 (-1.8 to -0.5) | 0.0001 |
| ISS, median (IQR) | 25 (17 - 29) | 25 (17 - 29) | 24 (17 - 26) | 25 (17 - 26) | 22 (17 - 26) | 24 (17 - 26) | -3 (-4 to -2) | <0.0001 |
| Head AIS 3+, n (%) | 1898 (63.6%) | 1959 (64.6%) | 6038 (67.2%) | 14724 (66.4%) | 19113 (62.5%) | 157542 (65.2%) | -1.1 (-2.9 to 0.7) | 0.224 |
| Spine AIS 3+, n (%) | 252 (8.4%) | 278 (9.2%) | 957 (10.7%) | 1961 (8.8%) | 2684 (8.8%) | 21830 (9%) | 0.3 (-0.7 to 1.4) | 0.544 |
| Face AIS 3+, n (%) | 70 (2.3%) | 110 (3.6%) | 79 (0.9%) | 123 (0.6%) | 166 (0.5%) | 2308 (1%) | -1.8 (-2.3 to -1.2) | <0.0001 |
| Thorax AIS 3+, n (%) | 1009 (33.8%) | 1011 (33.3%) | 2713 (30.2%) | 7305 (32.9%) | 10398 (34%) | 79616 (33%) | 0.2 (-1.6 to 1.9) | 0.847 |
| Abdomen AIS 3+, n (%) | 294 (9.9%) | 287 (9.5%) | 642 (7.1%) | 1578 (7.1%) | 2031 (6.6%) | 17718 (7.3%) | -3.2 (-4.3 to -2.1) | <0.0001 |
| Pelvis AIS 3+, n (%) | 323 (10.8%) | 433 (14.3%) | 923 (10.3%) | 1773 (8%) | 2850 (9.3%) | 22614 (9.4%) | -1.5 (-2.7 to -0.3) | 0.007 |
| Upper limb AIS 3+, n (%) | 16 (0.5%) | 17 (0.6%) | 86 (1%) | 196 (0.9%) | 285 (0.9%) | 2035 (0.8%) | 0.4 (0.1 to 0.7) | 0.029 |
| Lower limb AIS 3+, n (%) | 467 (15.7%) | 396 (13.1%) | 809 (9%) | 1676 (7.6%) | 2119 (6.9%) | 20480 (8.5%) | -8.7 (-10 to -7.4) | <0.001 |
| Polytrauma, n (%) | 1048 (35.1%) | 1081 (35.7%) | 2446 (27.2%) | 5680 (25.6%) | 7277 (23.8%) | 64549 (26.7%) | -11.3 (-13.1 to -9.6) | <0.0001 |
| **Pathways** |  |  |  |  |  |  |  |  |

|  | 2000 | 2005 | 2010 | 2015 | 2019 | Total 00 –19 | Magnitude of % change | P-value magnitude of % change* |
| --- | --- | --- | --- | --- | --- | --- | --- | --- |
| First Hospital MTC, n (%) | 1431 (48%) | 1588 (52.4%) | 4858 (54.1%) | 12940 (58.4%) | 16418 (53.7%) | 134659(55.8%) | 5.7 (3.8 to 7.6) | <0.0001 |
| MTC any time, n (%) | 1621 (54.3%) | 1683 (55.5%) | 5346 (59.5%) | 15003 (67.7%) | 19123 (62.5%) | 154082(63.8%) | 8.2 (6.3 to 10) | <0.0001 |
| **Interventions** |  |  |  |  |  |  |  |  |
| pre-hospital Doctor, 1st hospital, n (%) | 280 (13%) | 251 (12%) | 735 (11.5%) | 1947 (11.3%) | 2200 (8.8%) | 18917 (10.2%) | -4.1 (-5.6 to -2.7) | <0.0001 |
| Had CT, n (%) | 676 (22.7%) | 1136 (37.5%) | 6687 (74.4%) | 19106 (86.2%) | 27174 (88.9%) | 192393 79.7%) | 66.2 (64.7 to 67.7) | <0.0001 |
| Time to CT (mins), 1st hospital, median (IQR) | 87 (60 - 145) | 93 (60 - 152) | 81 (42 - 172) | 59 (27 - 155) | 77 (32 - 192) | 71 (31 - 169) | -10 (-14.4 to -5.6) | <0.0001 |
| Had Operation within 24 hrs, n (%) | 710 (23.8%) | 771 (25.4%) | 2076 (23.1%) | 3987 (18%) | 4853 (15.9%) | 45965 (19%) | -7.9 (-9.5 to -6.4) | <0.0001 |
| Time to surgery (hrs), median (IQR) | 3.1 (1.3 - 6.4) | 3.5 (1.5 - 7.3) | 4.4 (1.6 - 11.7) | 5.7 (2.2 - 15) | 6.6 (2.5 - 15.6) | 4.9 (2 - 13.4) | 3.5 (2.9 to 4.1) | <0.0001 |
| TXA given, direct admission, n (%) | 0 (0%) | 0 (0%) | 16 (0.3%) | 3146 (18.3%) | 4940 (19.8%) | 25922 (13.9%) | 19.6 (19.1 to 20.1) † | <0.0001† |
| TXA given for those given blood, direct admission, n (%) | 0 (0%) | 0 (0%) | 5 (1.4%) | 755 (91.8%) | 1189 (93.1%) | 6507 (74.5%) | 91.7(89.8 to 93.5) † | <0.0001† |
| Intubation, n (%) | 1499 (50.3%) | 1103 (36.4%) | 1897 (21.1%) | 3955 (17.8%) | 4243 (13.9%) | 48553 (20.1%) | -36.4 (-38.2 to -34.5) | <0.0001 |
| Critical care admission, n (%) | 1281 (42.9%) | 1468 (48.4%) | 3817 (42.5%) | 7053 (31.8%) | 8094 (26.5%) | 80732 (33.4%) | -16.5 (-18.3 to -14.6) | <0.0001 |
| Seen by Consultant in ED | 1031(34.6%) | 974(32.1%) | 3082(34.3%) | 11772(53.1%) | 16069(52.5%) | 32928(48.6%) | 18 (16.2 to 19.8) | <0.001<0.001 |
| **Outcomes** |  |  |  |  |  |  |  |  |

|  | **2000** | **2005** | **2010** | **2015** | **2019** | **Total 00 –19** | **Magnitude of % change** | **P-value magnitude of % change\*** |
| --- | --- | --- | --- | --- | --- | --- | --- | --- |
| LOS-days, median (IQR) | 8 (1 - 21) | 10 (4 - 22) | 9 (4 - 21) | 9 (4 - 19) | 8 (4 - 18) | 9 (4 - 19) | 0 | 0.999 |
| LOS- days CC, median (IQR) | 4 (2 - 10) | 5 (2 - 11) | 5 (2 - 11) | 4 (1 - 10) | 4 (1 - 9) | 4 (2 - 10) | 0 | 0.999 |
| Known Mortality Outcome, n(%) | 2380 (79.8%) | 2548 (84%) | 7416 (82.5%) | 18086 (81.6%) | 25909 (84.7%) | 198073 (82%) | 4.9 (3.4 to 6.4) | <0.0001 |
| Mortality, n (%) | 650 (27.3%) | 469 (18.4%) | 1157 (15.6%) | 2897 (16%) | 3748 (14.5%) | 31613 (16%) | -12.8 (-14.7 to -11) | <0.0001 |

(TARN- Trauma Audit and Research Network, CT - Computed Tomography Scan, MR – Magnetic Resonance Scan, yrs= years, GCS – Glasgow Coma Scale, SBP=Systolic Blood pressure, mmHg = millimetres of mercury, %o2sat = percentage oxygen saturation, bpm-beats per minute, AIS3+= Abbreviated injury Scale injury of at least 3 out of 6 severity, MTC= major trauma centre, TXA= Tranexamic Acid, LOS=length of stay, CC=critical care). \* Chi square for categorical variable and Bonnet-Price test for difference of median for continuous variables. † Comparison between 2010 and 2019.

**Figure 3:**
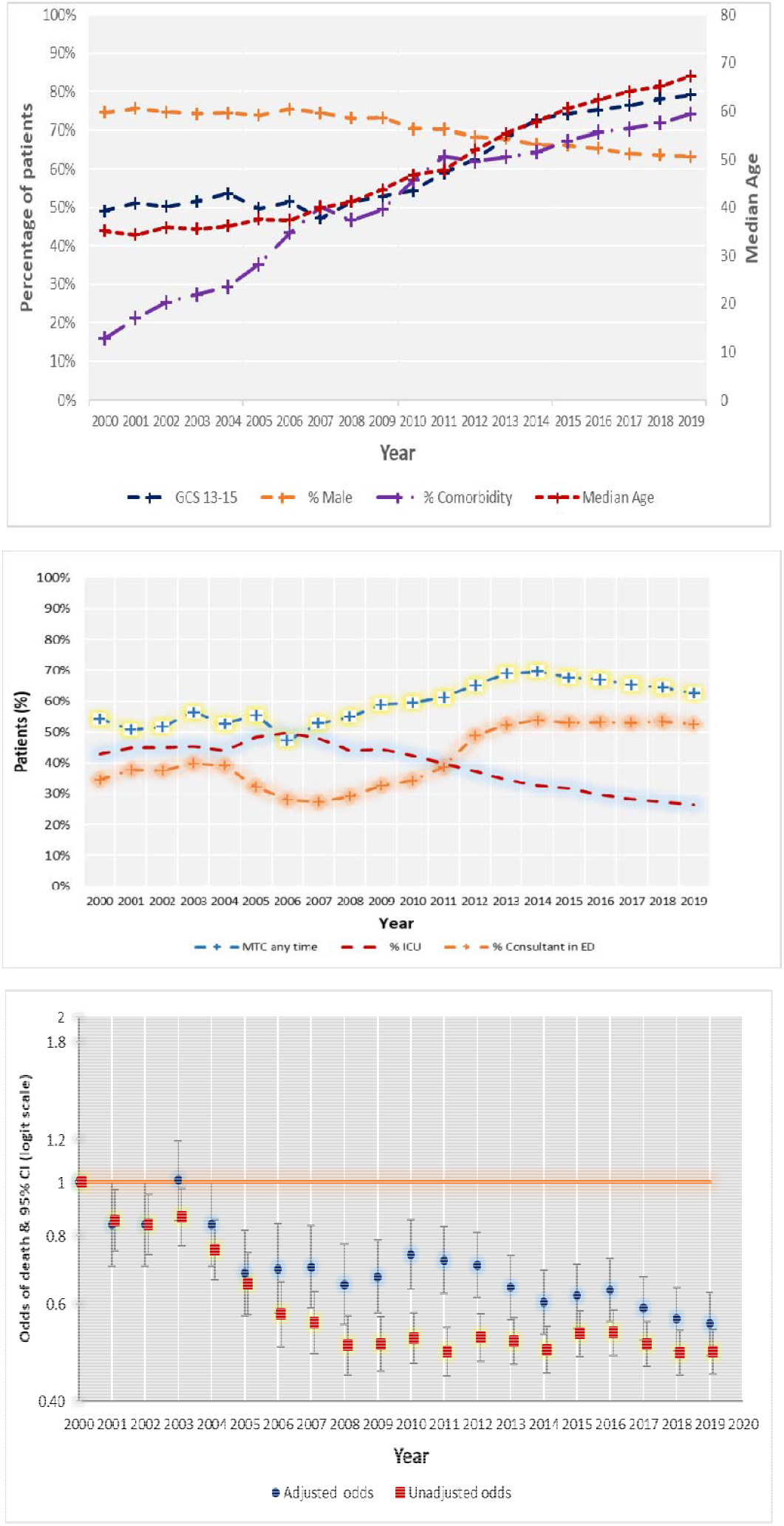
Contemporaneous annual Major Trauma patient characteristics (upper), care pathways (middle) and acute care mortality (lower) 2000 - 2019. Patient Characteristics; median age (years), percentage (i) GCS 13-15 at hospital arrival, (ii) male, (iii) with any comorbidity. Care processes - Percentage (i) managed at a Major Trauma Centre (MTC) at any time, (ii) admitted to critical care, (iii) managed by a Consultant in the Emergency Department. Mortality Annual adjusted and unadjusted odds of death compared to 2000 with 95% confidence limits. Computed Tomography imaging = CT, Magnetic Resonance Imaging =MR

Traumatic brain injury (TBI) and thoracic injuries have remained the dominant body region injuries across the whole cohort, being present in 65.2% and 33.0% of patients, respectively. There was a significant reduction in the proportion of patients with polytrauma of 11.3% (p<0.001). In terms of presenting physiology, patients in 2019 had less impairment in consciousness (proportion with GCS > 12 rose from 49% to 79%, Figure 3; chi squared test for trend p<0.001) compared to those presenting in 2000.

In terms of care pathways, over the study period initial presentation to a hospital classified (since 2012) as a Major Trauma Centre (MTC) increased from 48.0% (n=1,431) to 53.7% (n=16,418). Presentations to an MTC at any time in a patient’s care pathway rose from 54.3% (n=1,621) to 62.5% (n=19,123) (Table 2 and Figure 3; chi squared test for trend p<0.001). There was a significant decrease in the proportion of major trauma patients attended at scene by a pre-hospital doctor from 13% (n=280) to 8.8% (n=2200) and acute care endotracheal intubation decreased from 50.3% (n=1499) to 13.9% (n=4243). However, for both interventions the total recorded number of patients and numbers per hospital increased. An increasing proportion of patients received care from a consultant trauma team leader in the emergency department (52.5% versus 34.6%; Figure 3 - chi squared test for trend p<0.001).

Time from ED arrival to CT scan decreased from a median of 87 minutes in 2000 to 77 minutes in 2019. Between 2012 (when TXA became standard of care) and 2019, TXA administration on admission increased from 0.3% (n=16) to 19.8% (n=4940) and increased in those receiving blood products from 1.4% (n=5) to 93.1% (n=1189).

There was a decrease in the prevalence (but an increase in actual numbers per hospital) of major trauma patients requiring surgery within 24 hours from 23.8% (n=710) to 15.9% (n=4,853), with median hospital to operating suite arrival time interval increasing from 3.1 to 6.6 hours. Similarly, the proportion of patients requiring critical care admission decreased from 42.9% (n=1,281) to 26.5% (n=8,094; chi squared test for trend p<0.001) whilst actual numbers increased. Median length of stay overall and in ICU remained stable at 8 and 4 days, respectively. 18% of the study sample had unknown acute care mortality outcome arising from patients being transferred out alive of the first trauma receiving hospital but where no TARN data was sent by the final acute care hospital. In patients with known acute care mortality outcome unadjusted and risk adjusted mortality almost halved over the study period from 27.3% (n=650) in 2000 to 14.5% (n=3,748) in 2019, AOR 0.55 [95%CI 0.48 – 0.63] (Figure 3; chi squared tests for trend both p<0.001).

### Comparison of major trauma patients by causal energy transfer mechanism in 2019

By 2019 patients injured by low energy transfer (n=16,087) were older (median age 80 versus 47 years), less likely to be male (53.8% vs 73.8%), more likely to have pre-injury co-morbidity (90.4% versus 56.2%), less likely to have polytrauma (10.8% versus 38.2%) and more likely to have a traumatic brain injury visible on CT scan (74% versus 49.8%) than patients injured by high energy transfer (n=14,495) (Table 3). Although most high and low energy transfer patients presented with normal or only mildly impaired consciousness (GCS 13-15) the low energy cohort were less than half as likely to present with severe impairment of consciousness (GCS 3-8, 6% versus 15.3%; p<0.001) (Table 3). Overall, the anatomical severity of injury severity was greater in the high energy cohort (median ISS 25 versus 20 Table 3)

**Table 3:**
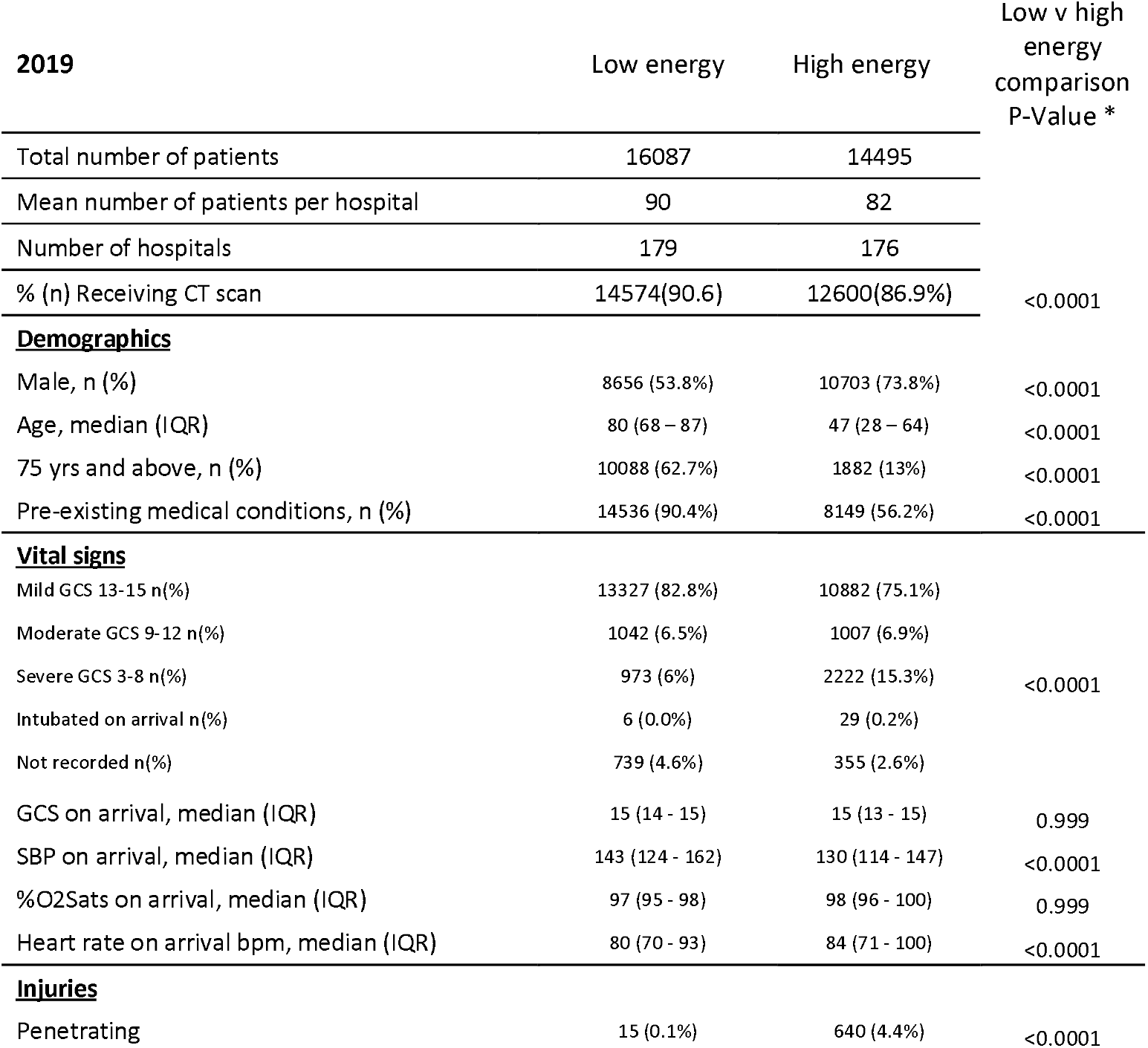
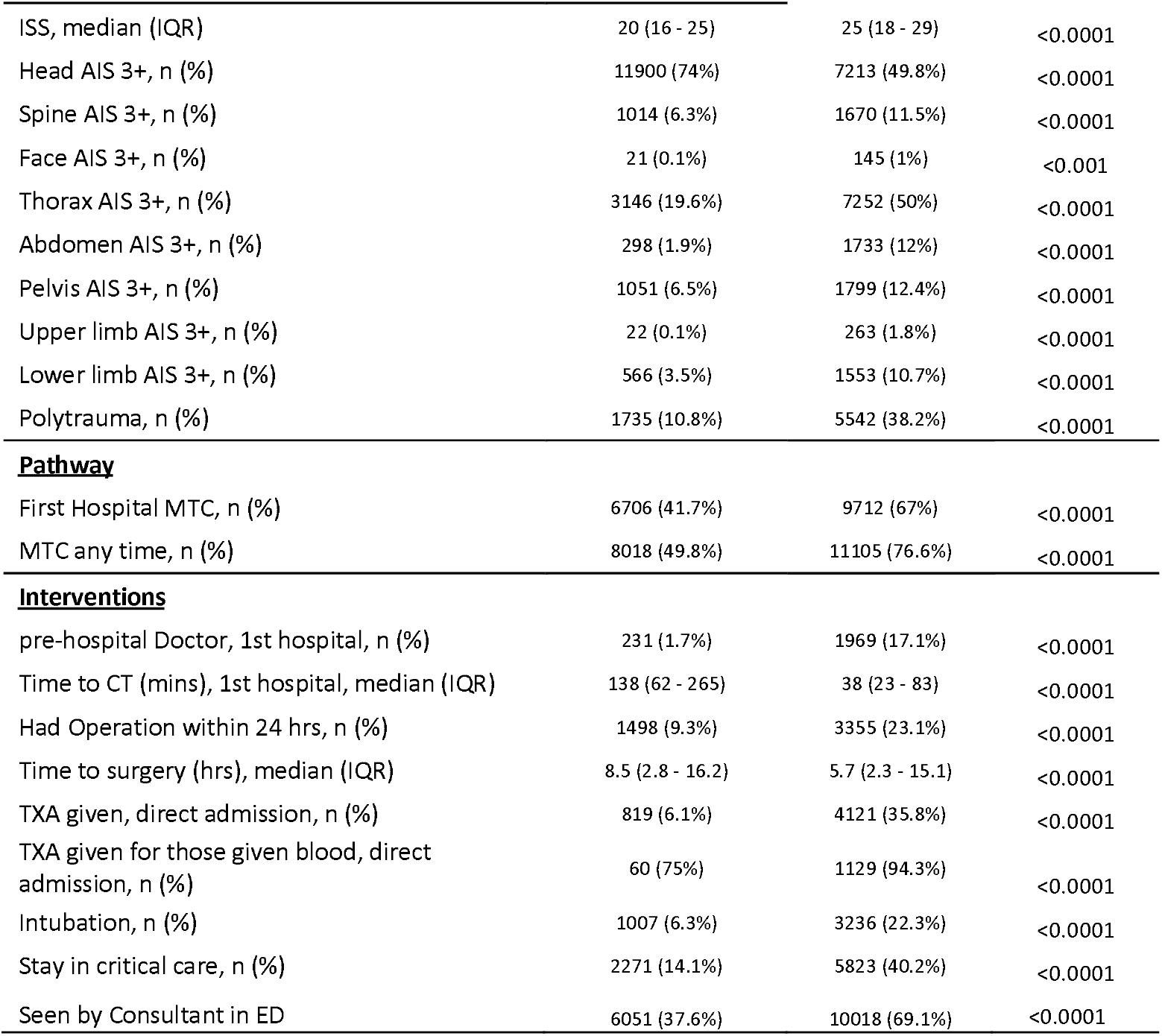
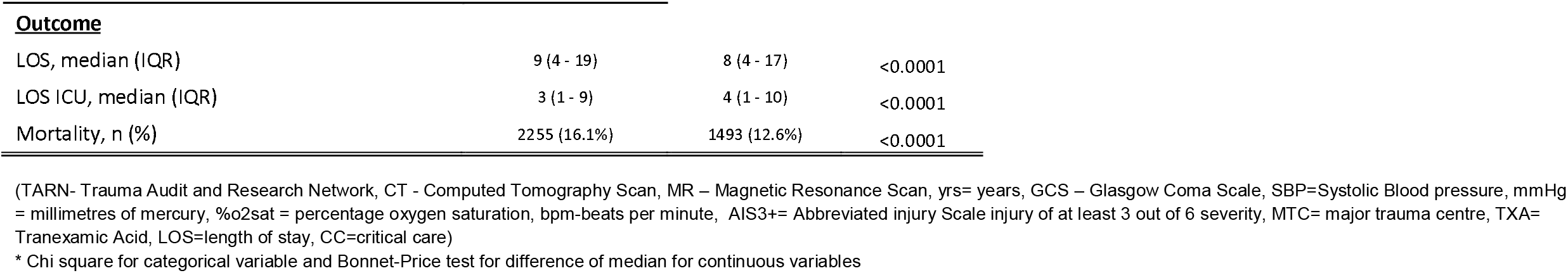
Comparison of major trauma patients injured by low versus high energy transfer in 2019 – all TARN Hospitals: Demographics; vital signs; injuries; pathway; interventions and outcomes.

| 2019 | Low energy | High energy | Low v high<br>energy<br>comparison<br>P-Value * |
| --- | --- | --- | --- |
| Total number of patients | 16087 | 14495 |  |
| Mean number of patients per hospital | 90 | 82 |  |
| Number of hospitals | 179 | 176 |  |
| % (n) Receiving CT scan | 14574(90.6) | 12600(86.9%) | <0.0001 |
| **<u>Demographics</u>** |  |  |  |
| Male, n (%) | 8656 (53.8%) | 10703 (73.8%) | <0.0001 |
| Age, median (IQR) | 80 (68 – 87) | 47 (28 – 64) | <0.0001 |
| 75 yrs and above, n (%) | 10088 (62.7%) | 1882 (13%) | <0.0001 |
| Pre-existing medical conditions, n (%) | 14536 (90.4%) | 8149 (56.2%) | <0.0001 |
| **<u>Vital signs</u>** |  |  |  |
| Mild GCS 13-15 n(%) | 13327 (82.8%) | 10882 (75.1%) |  |
| Moderate GCS 9-12 n(%) | 1042 (6.5%) | 1007 (6.9%) |  |
| Severe GCS 3-8 n(%) | 973 (6%) | 2222 (15.3%) | <0.0001 |
| Intubated on arrival n(%) | 6 (0.0%) | 29 (0.2%) |  |
| Not recorded n(%) | 739 (4.6%) | 355 (2.6%) |  |
| GCS on arrival, median (IQR) | 15 (14 - 15) | 15 (13 - 15) | 0.999 |
| SBP on arrival, median (IQR) | 143 (124 - 162) | 130 (114 - 147) | <0.0001 |
| %O2Sats on arrival, median (IQR) | 97 (95 - 98) | 98 (96 - 100) | 0.999 |
| Heart rate on arrival bpm, median (IQR) | 80 (70 - 93) | 84 (71 - 100) | <0.0001 |
| **<u>Injuries</u>** |  |  |  |
| Penetrating | 15 (0.1%) | 640 (4.4%) | <0.0001 |
| ISS, median (IQR) | 20 (16 - 25) | 25 (18 - 29) | <0.0001 |
| Head AIS 3+, n (%) | 11900 (74%) | 7213 (49.8%) | <0.0001 |
| Spine AIS 3+, n (%) | 1014 (6.3%) | 1670 (11.5%) | <0.0001 |
| Face AIS 3+, n (%) | 21 (0.1%) | 145 (1%) | <0.001 |
| Thorax AIS 3+, n (%) | 3146 (19.6%) | 7252 (50%) | <0.0001 |
| Abdomen AIS 3+, n (%) | 298 (1.9%) | 1733 (12%) | <0.0001 |
| Pelvis AIS 3+, n (%) | 1051 (6.5%) | 1799 (12.4%) | <0.0001 |
| Upper limb AIS 3+, n (%) | 22 (0.1%) | 263 (1.8%) | <0.0001 |
| Lower limb AIS 3+, n (%) | 566 (3.5%) | 1553 (10.7%) | <0.0001 |
| Polytrauma, n (%) | 1735 (10.8%) | 5542 (38.2%) | <0.0001 |
| **<u>Pathway</u>** |  |  |  |
| First Hospital MTC, n (%) | 6706 (41.7%) | 9712 (67%) | <0.0001 |
| MTC any time, n (%) | 8018 (49.8%) | 11105 (76.6%) | <0.0001 |
| **<u>Interventions</u>** |  |  |  |
| pre-hospital Doctor, 1st hospital, n (%) | 231 (1.7%) | 1969 (17.1%) | <0.0001 |
| Time to CT (mins), 1st hospital, median (IQR) | 138 (62 - 265) | 38 (23 - 83) | <0.0001 |
| Had Operation within 24 hrs, n (%) | 1498 (9.3%) | 3355 (23.1%) | <0.0001 |
| Time to surgery (hrs), median (IQR) | 8.5 (2.8 - 16.2) | 5.7 (2.3 - 15.1) | <0.0001 |
| TXA given, direct admission, n (%) | 819 (6.1%) | 4121 (35.8%) | <0.0001 |
| TXA given for those given blood, direct admission, n (%) | 60 (75%) | 1129 (94.3%) | <0.0001 |
| Intubation, n (%) | 1007 (6.3%) | 3236 (22.3%) | <0.0001 |
| Stay in critical care, n (%) | 2271 (14.1%) | 5823 (40.2%) | <0.0001 |
| Seen by Consultant in ED | 6051 (37.6%) | 10018 (69.1%) | <0.0001 |
| **<u>Outcome</u>** |  |  |  |
| LOS, median (IQR) | 9 (4 - 19) | 8 (4 - 17) | <0.0001 |
| LOS ICU, median (IQR) | 3 (1 - 9) | 4 (1 - 10) | <0.0001 |
| Mortality, n (%) | 2255 (16.1%) | 1493 (12.6%) | <0.0001 |

(TARN- Trauma Audit and Research Network, CT - Computed Tomography Scan, MR – Magnetic Resonance Scan, yrs= years, GCS – Glasgow Coma Scale, SBP=Systolic Blood pressure, mmHg = millimetres of mercury, %o2sat = percentage oxygen saturation, bpm=beats per minute, AIS3+= Abbreviated injury Scale injury of at least 3 out of 6 severity, MTC= major trauma centre, TXA= Tranexamic Acid, LOS=length of stay, CC=critical care)
\* Chi square for categorical variable and Bonnet-Price test for difference of median for continuous variables

Patients in the low energy transfer cohort were more likely to initially present to a Trauma Unit (TU) 58.3% versus 33% and less likely to receive MTC care at any time (49.8% versus 76.6%) than the high energy transfer cohort. They were seen less frequently by a doctor in the prehospital environment (1.7% versus 17%) or consultant in the emergency department (37.6% vs 69.1%). The low energy transfer cohort received TXA less frequently both on admission (6.1% vs 35.8%) and if administered blood products (75% vs 94.3%). Although rates of CT imaging were greater in the low energy cohort (90.6 versus 86.9%) they waited longer from arrival at hospital (median time to CT or MR 138 vs 38 mins), were less likely to require acute endotracheal intubation (6.3% vs 22.3%), be admitted to critical care (14.1% vs 40.2%), or require surgery within 24 hours (9.3% versus 23.1%); times from hospital arrival to surgery were also longer (8.5 versus 5.7 hours). The low energy cohort stayed on average 1 day (median nine versus eight days) longer in hospital but had shorter critical care stays (median three versus four days). Hospital mortality was 3.5% greater in the low energy cohort (16.1% [95%CI 15% - 17%] vs 12.6% [95%CI 12% - 13%]; p=<0.0001) (Table 3).

The sensitivity analysis in 19 consistently submitting hospitals revealed similar trends over time in major trauma patient casemix, care pathways and outcome, and in the comparison of patients injured by high and low energy transfer. As 85% of these hospitals are current Major Trauma Centres (as opposed to 15% nationally) the analysis is less well placed to assess trends in exposure to specialist care (suppl Tables 2 and 3, suppl Figures 2 and 3).

## Discussion

### Summary of findings

This study provides new insights into the changing aetiology and epidemiology of major trauma in England and Wales in the first two decades of the 21^st^ century. Falls from standing or less than two metres have become the predominant cause of major injury in 2019, quadrupling in prevalence since 2000. Our analysis suggests that the major drivers of this finding are increased use of CT/MR imaging, and improved trauma registry reporting systems, rather than population ageing. These changes are associated with a five-fold increase in the annual number of major trauma patients reported by each hospital.

Our findings suggest that changes in imaging practices and reporting to trauma registries – improving major trauma case ascertainment – explain the shift in the observed characteristics of major trauma patients in the NHS and internationally. Rather than a new epidemic of low energy trauma, it appears that improved access to imaging and trauma registry practices, together with the establishment of trauma networks in England and Wales, have unveiled a previously hidden burden of major trauma among patients who have fallen on the ground or from less than two metres. The observed changes cannot be explained by population demographic changes alone.

### Strengths and limitations of the study

This study is based on data from almost a quarter of a million patients with ISS >15 from Europe’s largest trauma registry and explores the potential drivers of the changing face of major trauma (3,18). TARN data is subject to multiple checks for accuracy and completeness supporting the internal validity of our findings. In addition, our analyses include data from the Office for National Statistics and comparisons to Hospital Episode Statistics (HES) which have allowed assessment of factors associated with the increasing low energy major trauma disease burden. Our multivariable models perform well using previously validated models (11).

Study limitations include changes in TARN membership over time; however, our sensitivity analyses in a core group of hospitals found the same increase in low energy prevalence with associated changes in casemix, care pathway and outcome over time (suppl Tables 1-3). A further limitation is the change in data fields collected over time. For example, rates of pre-injury anticoagulation are only available in later years, so could not be included in the analysis. The mortality analyses within this study are censored at acute care hospital discharge, but more recently (2014 onwards) TARN has collected 6-month post-injury patient reported functional and quality of life outcomes within MTCs, but these data could not be used in the longitudinal analysis. Furthermore, over the 20 years of 18% of patient mortality outcomes are missing, which is a limitation in the trends in mortality analyses which excluded these patients.

### Comparison with existing literature

Our findings are consistent with previous research, both in England and Wales(2,3,14) and in higher-income countries globally.(15–17) However, the associated potential drivers of the increasing low energy trauma disease burden have not previously been simultaneously examined.

Separating major trauma patients into high energy and low energy transfer cohorts helps to further define this emerging patient group, explaining the increasing age and comorbidity of the overall major trauma population, as well as highlighting crucial differences in the care pathways of these cohorts. A recent multicentre multinational registry based study from the EU funded Collaborative European Neurotrauma Effectiveness Research in TBI (CENTER-TBI) programme found similar differences in traumatic brain injury (TBI) care pathways for patients injured by low and high energy mechanisms and argued a specific approach is required to improve prevention and care for TBI patients injured through low falls(18).

The observation that in 2019 most of the low energy major trauma cohort (58.3% versus 33.0% of the high energy cohort) were first treated in a Trauma Unit and were less likely to receive consultant review suggests that current pre-hospital triage tools (which incorporate high energy mechanism of injury and deranged physiological parameters) are not sufficiently sensitive to detect the severity of their injuries. This is consistent with the findings of the CENTER TBI study. The challenge of accurately triaging older patients is well recognised(19–25), and the poor accuracy of all current methods was the finding of a recent systematic review(26). This may be explained by the different pathophysiology of traumatic brain injury in the two cohorts, highlighted by the greater likelihood of intracranial injury in the low energy cohort yet a higher presenting GCS (Table 3) (18).

### Interpretation of the study and implications

The reasons for improvements in major trauma case ascertainment are likely to be two fold; first in 2004 the first NICE head injury guideline recommended CT brain replace skull x-ray and observation as the primary diagnostic modality for patients with head injury and GCS 13-15 (mild traumatic brain injury – TBI), with a lower threshold for imaging patients aged 65 and over, the group more likely to sustain low energy major trauma (13). Figure 2 suggests a gradual adoption of these recommendations from 2004-2012 with an associated improved major trauma crude and risk adjusted mortality (Figure 3). The second reason may be that regional networks rewarded reporting to TARN through trauma best practice tariff payments; Figure 2 suggests this increased reporting to TARN significantly between 2012-17. This analysis suggests the burden of low energy major trauma will continue to increase as the population ages unless there are successful prevention initiatives, but the rate of increase is unlikely to replicate that seen in the first two decades of the 21^st^ century.

The lower rates of MTC admission at any time, surgical intervention, and admission to critical care in the low energy cohort are difficult to interpret. These findings may represent appropriate care as either patient factors (such as comorbidity, functional status, frailty and/ or ceiling of care decisions) preclude these interventions, or the pattern of injuries sustained from low energy trauma does not require them. However, there may also be systemic barriers to appropriate care for the low energy transfer group. Patient factors such as frailty (TARN commenced recording clinical frailty scores in mid-2019) and ceiling of care decisions (not currently collected) may be important for future care pathway analyses.

### Future research

These results highlight a pressing international need to examine how well the components of trauma networks and injury prevention initiatives serve the increasing volume of major trauma patients injured through falls from standing. Firstly, the drivers for differential care between the low and high energy transfer cohorts should be explored, particularly whether low energy trauma patients experience inappropriate barriers to accessing major trauma services. Secondly, the best care for the low energy transfer cohort needs to be defined so that integrated and inclusive trauma systems can optimise the use of both TU and MTC/highest level trauma centre resources. Thirdly, there is a need to understand how trauma units/lower-level trauma centres are managing major trauma patients both in the NHS and internationally. Our data suggest Trauma Units care for large volumes of patients injured by both high and low energy trauma (2). Lastly, there is a need to develop evidence-based interventions (building on successful multi-disciplinary approaches to hip fracture) to improve the mortality and functional outcomes of patients severely injured by low energy transfer mechanisms, particularly in those who have suffered TBI and thoracic injuries.

## Conclusion

Changes in major trauma patient imaging and reporting practices appear to have driven a marked increase in patient numbers over the last 20 years, revealing a previously hidden burden of major injury resulting from falls from standing or less than two metres. This cohort of patients is characterised by older age, more females, less abnormal physiology, but a high prevalence of traumatic brain and thoracic injuries. Despite the severity of their injuries, low energy transfer patients wait longer for investigation, are less likely to receive the highest-level major trauma care, to undergo surgery or be admitted to critical care and, in an unadjusted analysis, are more likely to die in hospital. Future research should recognise this distinct trauma population and investigate optimisation of trauma treatment networks and injury prevention strategies to mitigate the increasingly dominant burden of low energy transfer major trauma.

## Data Availability

Data cannot be shared as both: (i) the Data Sharing Agreements established between the TARN and member NHS Trusts, and (ii) the Section 251 HRA approval for analysis of anonymized TARN data specify the need for data access agreements with third parties. Proposals to access the study data, data dictionary, analytic code, and analysis scripts may be submitted online via www.tarn.ac.uk. Proposals are subject to review by the TARN Research committee. A Data Transfer Agreement is required, and all access must comply with TARN HRA approval.

## Contributorship Statement

FL, TC and AE designed the study. LW commented on data collection. OB performed statistical analysis. TS and MT interpreted the analysis and drafted the manuscript. All authors contributed to editing of the drafts and agreed with the final manuscript for submission. FL is the guarantor of the manuscript.

## Ethics Statement

TARN holds UK Health Research Authority Confidentiality Advisory Group approval for analysis of anonymised patient data - Section 251 NHS Act (2006).

## Public and Patient Involvement Statement

The TARN research committee approves all study proposals using TARN data and reports twice yearly to the TARN Board. TARN Board membership includes patient and public representation.

## Transparency Statement

The lead author (the manuscript’s guarantor) affirms that the manuscript is an honest, accurate, and transparent account of the study being reported; that no important aspects of the study have been omitted; and that any discrepancies from the study as originally planned (and, if relevant, registered) have been explained.

