## Supplementary material for "The changing major trauma disease burden from low falls in the first two decades of the 21st Century – a longitudinal analysis from the largest European Trauma Registry": Supp figures

**Supplemental materials**

Average marginal effects

|  | Effects on conditional mean of Low energy trauma |
| --- | --- |
| ONS 75+yrs | 0.175022 |
| % CT | 0.357606 |
| Case ascertainment | 0.342074 |

**Supplemental Figure 1:** Model showing average marginal effects of changes in annual (i) percentage of people over 75 years of age in England and Wales (ONS); (ii) percentage of TARN patients receiving CT/MRI imaging and (iii) percentage TARN v HES case ascertainment on changes in the relative annual prevalence of low energy transfer as causal injury mechanism – All TARN Hospitals 2000- 2019

Percentage of patients

Mean number of patients /hospital/year

| Year | Age 75+ | Had CT | Low Energy | ONS 75+ | Case ascertmnt |
| --- | --- | --- | --- | --- | --- |
| 2000 | 8.52515% | 20.6309% | 15.26% | 7.50827% |  |
| 2001 | 7.30337% | 24.2509% | 11.3296% | 7.57102% |  |
| 2002 | 8.74786% | 23.3276% | 11.235% | 7.60086% |  |
| 2003 | 7.04981% | 26.8966% | 10.8046% | 7.60708% |  |
| 2004 | 8.48% | 28.96% | 13.44% | 7.61817% |  |
| 2005 | 8.48921% | 29.0647% | 14.4604% | 7.63327% |  |
| 2006 | 7.4856% | 37.524% | 15.739% | 7.67451% |  |
| 2007 | 9.26766% | 51.523% | 14.9708% | 7.7056% |  |
| 2008 | 11.4487% | 58.9257% | 17.9056% | 7.71442% | 50.6% |
| 2009 | 13.0688% | 65.7721% | 21.198% | 7.72916% | 61.6% |
| 2010 | 14.381% | 68.6032% | 22.9206% | 7.78087% | 67.2% |
| 2011 | 17.2992% | 74.505% | 27.8053% | 7.83535% | 71.9% |
| 2012 | 20.0779% | 80.1655% | 30.3967% | 7.91927% | 83.7% |
| 2013 | 22.4969% | 83.1222% | 32.1823% | 7.99453% | 88.35% |
| 2014 | 23.8037% | 85.4881% | 34.4702% | 8.1018% | 93.2% |
| 2015 | 25.7576% | 85.1994% | 36.0925% | 8.12725% | 95.3% |
| 2016 | 27.9915% | 85.1215% | 37.6943% | 8.13874% | 96.5% |
| 2017 | 28.7115% | 84.7706% | 42.9322% | 8.20838% | 96.5% |
| 2018 | 28.2557% | 84.6871% | 39.3342% | 8.32995% | 96.1% |
| 2019 | 31.117% | 85.7634% | 43.1328% |  | 97.25% |

|  | Mean number/hospital/year |
| --- | --- |
| 2000 | 61.7368 |
| 2001 | 56.2105 |
| 2002 | 61.3684 |
| 2003 | 68.6842 |
| 2004 | 65.7895 |
| 2005 | 73.1579 |
| 2006 | 54.8421 |
| 2007 | 81.2105 |
| 2008 | 97 |
| 2009 | 135.316 |
| 2010 | 165.789 |
| 2011 | 191.368 |
| 2012 | 216.263 |
| 2013 | 257.579 |
| 2014 | 302.474 |
| 2015 | 330 |
| 2016 | 348.789 |
| 2017 | 370.474 |
| 2018 | 395.263 |
| 2019 | 398.158 |

Supplemental figure 2: For consistently submitting hospitals (n=19) contemporaneous annual (2000-2019) percentage of TARN ISS>15 patients injured through low energy transfer (primary outcome) and factors assessed for temporal association; percentage of TARN (i) aged 75 years and over (ii) receiving CT or MR scanning; percentage of population of England and Wales aged 75 years and over, mean number of ISS>15 patients reported per hospital to TARN, percentage TARN case ascertainment (denominator = TARN eligibility in Hospital Episode Statistics 2008-2019). (TARN- Trauma Audit and Research Network, CT - Computed Tomography Scan, MR – Magnetic Resonance Scan, ONS – Office of National Statistics, HES – Hospital Episode Statistics)

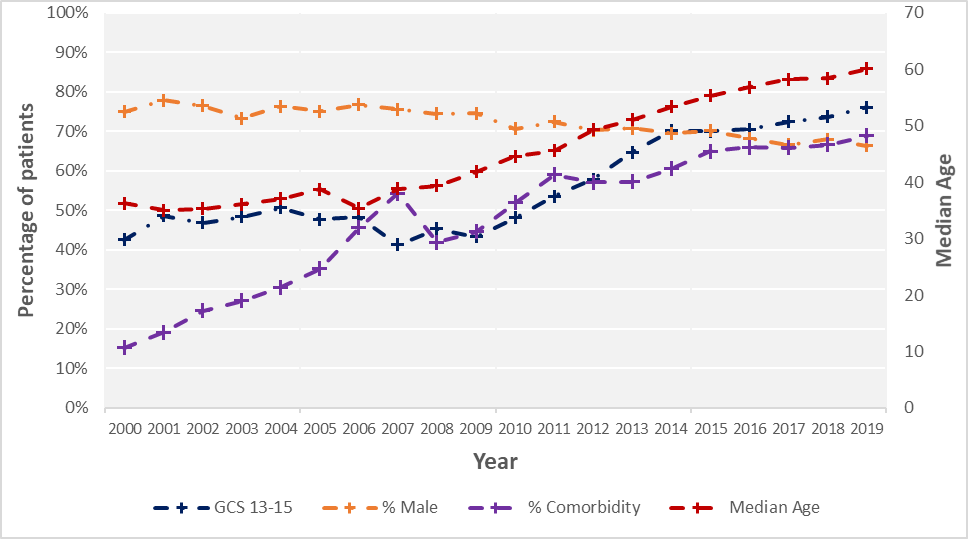

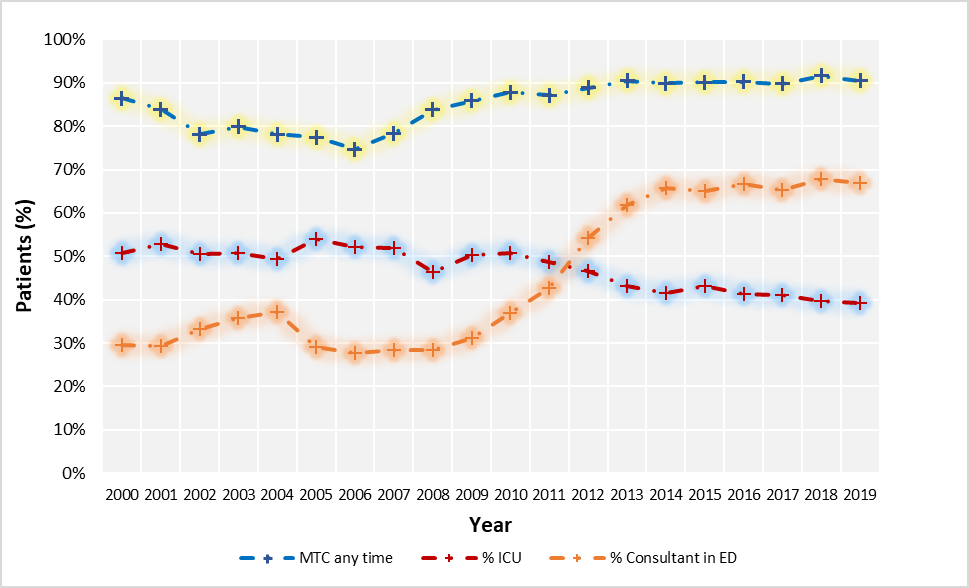

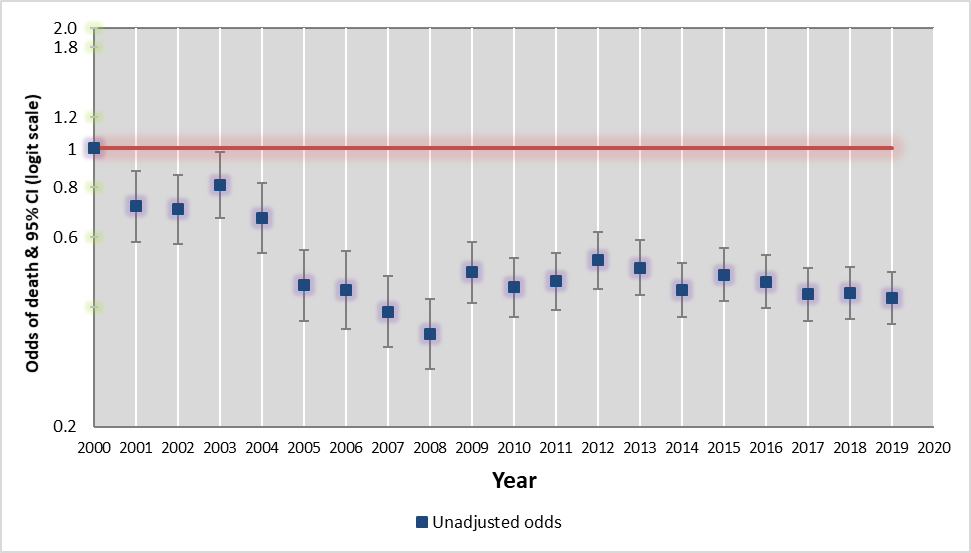

**Supplemental figure 3:** For consistently submitting hospitals (n=19) contemporaneous annual Major Trauma patient characteristics (upper), care pathways (middle), crude mortality (lower) 2000 - 2019. Patient Characteristics: median age (years), percentage (i) GCS 13-15 at presentation, (ii) male, (iii) with any comorbidity. Care processes - Percentage (i) managed at a Major Trauma Centre (MTC) at any time, (ii) admitted to critical care, (iii) percentage managed by consultant in the Emergency Department

**Supplemental table 1** Annual prevalence of patients injured by low energy major trauma (primary outcome) and factors assessed for association in 19 hospitals consistently submitting to TARN in 2000, 2005, 2010, 2015, 2019.

|  | **2000** | **2005** | **2010** | **2015** | **2019** |
| --- | --- | --- | --- | --- | --- |
| Patients with ISS > 15 reported to TARN – N | 1173 | 1390 | 3150 | 6270 | 7565 |
| TARN member hospitals in calendar year- N | 19 | 19 | 19 | 19 | 19 |
| Mean number of patients per hospital with ISS >15 reported to TARN | 62 | 73 | 166 | 330 | 398 |
| TARN patients with ISS >15 receiving CT+/or MR imaging; (%) | 20.63% | 29.06% | 68.60% | 85.20% | 85.76% |
| TARN v HES % case ascertainment | n/a | n/a | 73.6% | 87.2% | 93.4% |
| England and Wales population aged over 75 years; N (%) | 3914824 (7.5%) | 4089552 (7.6%) | 4333354 (7.8%) | 4704493 8.1%) | 5078598 (8.5%) |
| TARN patients with ISS >15 aged over 75 years; N (%) | 100(8.5%) | 118(8.5%) | 453(14.4%) | 1615(25.8%) | 2354(31.1%) |
| Prevalence of low energy transfer as causal mechanism in TARN ISS > 15; N (%) | 179(15.3%) | 201(14.5%) | 722(22.9%) | 2263(36.1%) | 3263(43.1%) |

(TARN- Trauma Audit and Research Network, CT - Computed Tomography Scan, MR – Magnetic Resonance Scan, HES – Hospital Episode Statistics, n/a= not available)

**Supplemental Table 2**: Annual Major Trauma Patient Cohort Characteristics: case mix, care pathway and acute care outcomes in 2000, 2005, 2010, 2015, 2019: 19 hospitals consistently submitting to TARN

|  | 2000 | 2005 | 2010 | 2015 | 2019 | Overall, 2000 - 2019 | Magnitude of % change | P-Value magnitude of % change |
| --- | --- | --- | --- | --- | --- | --- | --- | --- |
| **Demographics** | | | | | | | | |
| Male, n (%) | 880(75%) | 1043(75%) | 2226(70.7%) | 4402(70.2%) | 5022(66.4%) | 49765(70.2%) | -9 ( -11 to -6) | <0.0001 |
| Age, median (IQR) | 36.3 (23.2 - 54.2) | 38.8 (23.3 - 56.3) | 44.6 (26 - 63.2) | 55.3 (33.3 - 75.6) | 60.1 (37 - 79) | 50.9 (29.9 - 72.5) | 23.8 (21.9 to 25.6) | <0.0001 |
| 0 - 24 yrs, n (%) | 336(28.6%) | 395(28.4%) | 723(23%) | 973(15.5%) | 954(12.6%) | 13038(18.4%) | -16 (-18.7 to -13) | <0.0001 |
| 25 - 49 yrs, n (%) | 473(40.3%) | 536(38.6%) | 1113(35.3%) | 1733(27.6%) | 1890(25%) | 21548(30.4%) | -15.3 ( -18.3 to -12.4) | <0.0001 |
| 50 - 74 yrs, n (%) | 264(22.5%) | 341(24.5%) | 861(27.3%) | 1949(31.1%) | 2367(31.3%) | 20605(29.1%) | 8.8 (6.2 to 11.4) | <0.0001 |
| 75 yrs and above, n (%) | 100(8.5%) | 118(8.5%) | 453(14.4%) | 1615(25.8%) | 2354(31.1%) | 15707(22.2%) | 22.6 (20.7 to 24.5) | <0.0001 |
| Pre-existing medical conditions, n (%) | 179(15.3%) | 490(35.3%) | 1637(52%) | 4071(64.9%) | 5211(68.9%) | 40833(57.6%) | 53.6 (51.3 to 55.9) | <0.0001 |
| **Mechanism** | | | | | | | | |
| High energy trauma | 994(84.7%) | 1189(85.5%) | 2428(77.1%) | 4007(63.9%) | 4302(56.9%) | 48149(67.9%) | -27.9 (-30.2 to -25.5) | <0.0001 |
| Low energy trauma | 179(15.3%) | 201(14.5%) | 722(22.9%) | 2263(36.1%) | 3263(43.1%) | 22749(32.1%) | 27.9(25.5 to 30.2) | <0.0001 |
| **Vital signs** | | | | | | | | |
| Mild GCS 13-15 n(%) | 499 (42.5%) | 664 (47.8%) | 1515 (48.1%) | 4392 (70.1%) | 5753 (76.1%) | 24902(53.6%) | 33.5 (33.5 to 36.5) | <0.0001 |
| Moderate GCS 9-12 n (%) | 147 (12.5%) | 118 (8.5%) | 288 (9.1%) | 586 (9.4%) | 613 (8.1%) | 5381(11.6%) | -4.4 (-6.4 to --2.4) | <0.0001 |
| Severe 3-8 n (%) | 421 (35.9%) | 329 (23.7%) | 480 (15.2%) | 990 (15.8%) | 950 (12.6%) | 10231(22%) | -23.3 (-26.2 to -20.5) | <0.0001 |
| GCS Not recorded n (%) | 106 (9%) | 279 (20.1%) | 867 (27.5%) | 302 (4.8%) | 249 (3.3%) | 5988(12.9%) | -5.7 (-7.4 to - 4) | <0.0001 |
| GCS on arrival, median (IQR) | 12(5-15) | 14(7-15) | 14(10-15) | 15(12-15) | 15(13-15) | 15(11-15) | 3 (2.5 to 3.5) | <0.0001 |
| SBP on arrival, median (IQR) | 130(115-150) | 130(113-150) | 132(115-150) | 132(116-151) | 132(115-150) | 132(115-150) | 2 (0.6 to 3.4) | 0.005 |
| O2Sats on arrival, median (IQR) | 99(96-100) | 99(97-100) | 98(96-100) | 98(96-100) | 98(96-100) | 98(96-100) | -1 (-1.49 to -0.5) | 0.0001 |
| Heart rate on arrival, median (IQR) | 88(75-106) | 88(74-105) | 88(74-106) | 84(71-100) | 83(71-97) | 85(72-100) | -5 (-7.2 to -2.8) | <0.0001 |
| **Injuries** | | | | | | | | |
| Penetrating, n (%) | 41(3.5%) | 70(5%) | 111(3.5%) | 162(2.6%) | 212(2.8%) | 2236(3.2%) | -0.69 ( -1.9 to 0.4) | 0.188 |
| ISS, median (IQR) | 25(18-29) | 25(17-29) | 25(17-29) | 25(17-29) | 25(17-27) | 25(17-29) |  |  |
| Head AIS 3+, n (%) | 843(71.9%) | 944(67.9%) | 2068(65.7%) | 4137(66%) | 4658(61.6%) | 46502(65.6%) | -10.3 ( -13.1 to -7.5) | <0.0001 |
| Spine AIS 3+, n (%) | 84(7.2%) | 143(10.3%) | 333(10.6%) | 616(9.8%) | 780(10.3%) | 6989(9.9%) | 3.1 (1.5 to 4.8) | 0.0008 |
| Face AIS 3+, n (%) | 32(2.7%) | 58(4.2%) | 30(1%) | 45(0.7%) | 66(0.9%) | 903(1.3%) | -1.8 (-2.8 to -0.9) | <0.0001 |
| Thorax AIS 3+, n (%) | 378(32.2%) | 442(31.8%) | 1045(33.2%) | 2245(35.8%) | 2803(37.1%) | 24920(35.1%) | 4.8 (1.9 to 7.7) | 0.002 |
| Abdomen AIS 3+, n (%) | 98(8.4%) | 123(8.8%) | 256(8.1%) | 467(7.4%) | 643(8.5%) | 5659(8.0%) | 1.4 (-1.5 to 1.8) | 0.868 |
| Pelvis AIS 3+, n (%) | 121(10.3%) | 169(12.2%) | 419(13.3%) | 585(9.3%) | 827(10.9%) | 7446(10.5%) | 6.2 (-13 to 2.5) | 0.528 |
| Upper limb AIS 3+, n (%) | 7(0.6%) | 11(0.8%) | 36(1.1%) | 62(1.0%) | 93(1.2%) | 682(1.0%) | 0.6 (0.1 to 1,1) | 0.581 |
| Lower limb AIS 3+, n (%) | 163(13.9%) | 174(12.5%) | 320(10.2%) | 535(8.5%) | 602(8.0%) | 6590(9.3%) | -5.9 (-8 to -3.9) | <0.0001 |
| Polytrauma, n (%) | 414(35.3%) | 492(35.4%) | 959(30.4%) | 1848(29.5%) | 2245(29.7%) | 21741(30.7%) | -5.6(-8.5 to -2.7) | 0.0001 |
| **Pathway** | | | | | | | | |
| First Hospital MTC, n (%) | 1002(85.4%) | 1058(76.1%) | 2687(85.3%) | 5464(87.1%) | 6634(87.7%) | 60604(85.5%) | 2.3 (0.1 to 4.4) | 0.029 |
| MTC any time, n (%) | 1014(86.4%) | 1076(77.4%) | 2768(87.9%) | 5649(90.1%) | 6842(90.4%) | 62507(88.2%) | 4 (1.9 to 6.1) | <0.0001 |
| **Interventions** | | | | | | | | |
| pre-hospital Doctor, 1st hospital, n (%) | 173(23.8%) | 154(18.3%) | 457(23.7%) | 831(19%) | 660(12.2%) | 7813(16.3%) | -1.1 (-15 to -8.3) | <0.0001 |
| Had CT, n (%) | 242(20.6%) | 404(29.1%) | 2161(68.6%) | 5342(85.2%) | 6488(85.8%) | 52796(74.5%) | 65 (62.7 to 67.6) | <0.0001 |
| Time to CT (mins), 1st hospital, median (IQR) | 77(55-124) | 89.5(56-140.5) | 61(33-139) | 39(23-98) | 45(25-128) | 49(26-120) |  |  |
| Had Operation within 24 hrs, n (%) | 316(26.9%) | 413(29.7%) | 964(30.6%) | 1405(22.4%) | 1804(23.8%) | 18148(25.6%) | -3.1 (-5.8 to -0.4) | 0.0215 |
| Time to surgery (hrs), median (IQR) | 2.67(1.13-5.68) | 3.09(1.16-6.52) | 4.50(1.49-12.38) | 5.50(2.18-14.57) | 6.26(2.28-14.78) | 4.67(1.78-12.90) | 3.6 (2.8 to 4.4) | <0.0001 |
| TXA given, direct admission, n (%) | 0(0) | 0(0) | 8(0.4%) | 1063(24.4%) | 1475(27.3%) | 8249(17.2%) | 26.8 (26 to28) † | <0.0001† |
| TXA given for those given blood, direct admission, n (%) | 0(0%) | 0(0%) | 2(1.3%) | 215(93.9%) | 368(92.7%) | 1807(71.1%) | 91.3 (88.2 to 94) † | <0.0001 † |
| Intubation, n (%) | 649(55.3%) | 510(36.7%) | 713(22.6%) | 1419(22.6%) | 1367(18.1%) | 17434(24.6%) | -37 ( -40 to -34) | <0.0001 |
| StayCC Critical care, n (%) | 595(50.7%) | 750(54%) | 1595(50.6%) | 2709(43.2%) | 2969(39.2%) | 31319(44.2%) | -11.5 (-14.5 to -8.4) | <0.0001 |
| Seen by Consultant in ED | 346(29.5%) | 403(29%) | 1161(36.9%) | 4085(65.2%) | 5068(67%) | 11063(56.6%) | 37.5 (34.7 to 40.3) | <0.0001 |
| **Outcomes** | | | | | | | | |
| LOS, median (IQR) | 9(3-20) | 11(5-25) | 11(5-23) | 10(5-21) | 10(5-20) | 10(5-22) | 1 (0.5 to 1.5) | 0.0001 |
| LOS ICU, median (IQR) | 4(2-10) | 6(2-13) | 5(2-12) | 4(1-11) | 4(1-10) | 4(2-11) | 0 (-0.5 to 0.5) | 0.999 |
| Known Mortality Outcome, n(%) | 1027 (87.6%) | 1211 (87.1%) | 2571 (81.6%) | 5237 (83.5%) | 6653 (87.9%) | 59701 (84.2%) | 0.4(-1.6 to 2.4) | 0.702 |
| Mortality, n (%) | 299(29.1%) | 190(15.7%) | 399(15.5%) | 864(16.5%) | 982(14.8%) | 9763(16.4%) | -14.7 (-17.2 to -11.4) | <0.0001 |

(TARN- Trauma Audit and Research Network, CT - Computed Tomography Scan, MR – Magnetic Resonance Scan, yrs= years, GCS – Glasgow Coma Scale, SBP=Systolic Blood pressure, mmHs = millimetres of mercury, %o2sat = percentage oxygen saturation, bpm-beats per minute, AIS3+= Abbreviated injury Scale injury of at least 3 out of 6 severity, MTC= major trauma centre, TXA= Tranexamic Acid, LOS=length of stay, CC=critical care). † Comparison between 2010 and 2019.

**Supplemental Table 3**: Comparison of major trauma patients injured by low versus high energy transfer in 2019 receiving care at 19 TARN Hospitals (consistently submitting data from 2000): Demographics; vital signs; injuries; pathway; interventions and outcomes.

| **2019** | Low energy | High energy |
| --- | --- | --- |
| Total number of patients | 2363 | 4302 |
| Mean number of patients per hospital | 124 | 226 |
| Number of hospitals | 19 | 19 |
| % Receiving CT scan | 85.8% | 85.7% |
| **Demographics** |  |  |
| Male, n (%) | 1823(55.9%) | 3199(74.4%) |
| Age, median (IQR) | 78.2(64.7 - 86) | 44.5(27.5 - 62) |
| 75 yrs and above, n (%) | 480(11.2%) | 1874(57.4%) |
| Pre-existing medical conditions, n (%) | 2859(87.6%) | 2352(54.7%) |
| **Vital signs** |  |  |
| Mild GCS 13-15 n(%) | 2642 (81%) | 3111 (72.3%) |
| Moderate GCS 9-12 n(%) | 255 (7.8%) | 358 (8.3%) |
| Severe GCS 3-8 n(%) | 243 (7.4%) | 707 (16.4%) |
| Intubated on arrival n(%) | 3 (0.1%) | 10 (0.2%) |
| Not recorded n(%) | 120 (3.7%) | 116 (2.7%) |
| GCS on arrival, median (IQR) | 15(14-15) | 15(12-15) |
| SBP on arrival, median (IQR) | 140(121-159) | 127(113-143) |
| O2Sat on arrival, median (IQR) | 97(95-99) | 98(96-100) |
| Heart rate on arrival, median (IQR) | 80(69-94) | 84(72-100) |
| **Injuries** |  |  |
| Penetrating | 4(0.1%) | 208(4.8%) |
| ISS, median (IQR) | 25(16-25) | 25(19-30) |
| Head AIS 3+, n (%) | 2412(73.9%) | 2246(52.2%) |
| Spine AIS 3+, n (%) | 250(7.7%) | 530(12.3%) |
| Face AIS 3+, n (%) | 7(0.2%) | 59(1.4%) |
| Thorax AIS 3+, n (%) | 641(19.6%) | 2162(50.3%) |
| Abdomen AIS 3+, n (%) | 72(2.2%) | 571(13.3%) |
| Pelvis AIS 3+, n (%) | 239(7.3%) | 588(13.7%) |
| Upper limb AIS 3+, n (%) | 5(0.2%) | 88(2.0%) |
| Lower limb AIS 3+, n (%) | 100(3.1%) | 502(11.7%) |
| Polytrauma, n (%) | 420(12.9%) | 1825(42.4%) |
| **Pathway** |  |  |
| First Hospital MTC, n (%) | 2652(81.3%) | 3982(92.6%) |
| MTC any time, n (%) | 2754(84.4%) | 4088(95%) |
| **Interventions** |  |  |
| pre-hospital Doctor, 1st hospital, n (%) | 63(2.8%) | 597(18.9%) |
| Had CT, n (%) | 2801(85.8%) | 3687(85.7%) |
| Time to CT (mins), 1st hospital, median (IQR) | 111(43-260) | 31(21-56) |
| Had Operation within 24 hrs, n (%) | 526(16.1%) | 1278(29.7%) |
| Time to surgery (hrs), median (IQR) | 8.75(2.55-16.18) | 5.58(2.22-14.23) |
| TXA given, direct admission, n (%) | 167(7.4%) | 1308(41.4%) |
| TXA given for those given blood, direct admission, n (%) | 12(63.2%) | 356(94.2%) |
| Intubation, n (%) | 293(9%) | 1074(25%) |
| Stay in critical care, n (%) | 727(22.3%) | 2242(52.1%) |
| Seen by Consultant in ED, n (%) | 1661(50.9%) | 3407(79.2%) |
| **Outcome** |  |  |
| LOS, median (IQR) | 10(5-20) | 10(5-19) |
| LOS ICU, median (IQR) | 4(1-9) | 4(2-10) |
| Mortality, n (%) | 493(17.2%) | 489(12.9%) |

(TARN- Trauma Audit and Research Network, CT - Computed Tomography Scan, MR – Magnetic Resonance Scan, yrs= years, GCS – Glasgow Coma Scale, SBP=Systolic Blood pressure, mmHs = millimetres of mercury, %o2sat = percentage oxygen saturation, bpm-beats per minute, AIS3+= Abbreviated injury Scale injury of at least 3 out of 6 severity, MTC= major trauma centre, TXA= Tranexamic Acid, LOS=length of stay, CC=critical care)
